# Computational modeling of seizure onset patterns to underpin their underlying mechanisms

**DOI:** 10.1101/2023.02.16.23286033

**Authors:** Leila Abrishami Shokooh, Frédéric Lesage, Dang Khoa Nguyen

**Affiliations:** École Polytechnique de Montréal, Université de Montréal, C.P. 6079, succ. Centre-Ville, Montreal, QC H3C 3A7, Canada; Centre de Recherche du Centre Hospitalier de l’Université de Montreal (CHUM), Montreal, QC H2X 0A9, Canada; Research Centre, Montreal Heart Institute, Montreal, QC H1T 1C8, Canada; Neurology Division, Centre Hospitalier de l’Université de Montréal (CHUM), 1000 Saint-Denis, Montreal, QC H2X 0C1, Canada

## Abstract

In the study of epilepsy, it is of crucial importance to understand the transition from interictal into ictal activities (ictogenesis). Different mechanisms have been suggested for the generation of ictal activity; yet, it remains unclear whether different physiological mechanisms underly different seizure onset patterns. Herein, by implementing a computational model that takes into account some of the most relevant physiological events (e.g., depolarization block, collapse, and recovery of inhibitory activities) and different scenarios of imbalanced excitatory-inhibitory activities, we explored if seizures with different onset patterns stem from different underlying mechanisms. Our model revealed that depending on the excitation level, seizures could be generated due to both enhancement and collapse of inhibition for specific range of parameters. Successfully reproducing some of the commonly observed seizure onset patterns, our findings indicated that different onset patterns can arise from different underlying mechanisms.

**Significance Statement:** Various seizure onset patterns have been reported; however, it yet remains unknown whether seizures with distinct onset patterns originate from different underlying mechanisms. The common belief on seizure generation focuses on the imbalance between synaptic excitation and inhibition which has led to the identification of distinct and, in some cases, even contradictory mechanisms for seizure initiation. In this study, by incorporating some of these various physiological mechanisms in a unified framework, we reproduced some commonly observed seizure onset patterns. Our results suggest the existence of different mechanisms responsible for the generation of seizures with distinct onset patterns which can enhance our understanding of seizure generation mechanisms with significant implications on developing therapeutic measures in seizure control.

## Introduction

Epilepsy, second only to stroke, is one of the most common chronic neurological disorders. It affects 50 million people worldwide [1] and is characterized by recurrent spontaneous seizures, i.e., sudden transient behavioral disturbances due to abnormal excessive neural discharges [2]. About 30% of patients have drug-resistant epilepsy [3–5] which could be due to the diverse network and cellular mechanisms responsible for seizure generation [6].

Variable electrophysiological features have been reported for seizures in intracranial EEG studies which has resulted in the definition of specific patterns for seizure onset [7,8]. Some onset patterns have been associated with the activity of specific brain regions [9–11] and synaptic mechanisms [12,13]. For instance, using an in vitro optogenetic experimental setting, it has been reported that seizures with Low Voltage Fast Activity (LVFA) onset are generated due to the involvement of hyperpolarizing inhibitory events while the generation of seizures with hypersynchronous onset depends on enhanced excitation [12,13]. These findings raise the question as to whether distinct onset patterns of ictal activity arise from fundamentally different mechanisms.

The mechanisms underlying seizure generation involve the interaction of many complex cellular and synaptic components [14,15] which could govern the emergence of distinct iEEG ictal patterns [16–18]. Studying the imbalance between synaptic excitation and inhibition has been the main focus of the research on ictogenesis [13,19–24]. It was commonly believed that abnormally enhanced glutamatergic excitation and collapse of GABAergic inhibition were pro-epileptic and the main driver for seizure generation, whereas the increased GABAergic activity was viewed to protect against seizure generation [25–28]. However, this notion might require reconsideration in light of more recent clinical and experimental data that have offered evidence for the critical role of the GABAergic network in focal seizures. More specifically, some studies have reported enhancement of inhibitory activity before and at the very onset of ictal activity [29–33], casting doubt on the suggested role for the collapse of inhibition in seizure generation. In summary, distinct and even contradictory mechanisms have been suggested to underly seizure initiation.

Incorporating these distinct mechanisms in a unified framework would enable us to compare the impact of these different mechanisms in seizure generation and would aid in providing an insight into whether different underlying mechanisms could lead to seizures with different onset patterns.

To investigate the underlying mechanisms for generation of some of the most common patterns of ictal activity, we constructed a neural mass model and, by updating, expanding, and exploring a well-established framework [34,35], we generated different patterns of seizure generation. More specifically, we considered different scenarios of disturbed balance between excitatory and inhibitory activities and identified a range of parameters producing distinct onset patterns providing interpretation in terms of underlying physiological mechanisms. The model successfully reproduced a number of commonly observed onset patterns including Low Voltage Fast Activity (LVFA), spindles of alpha/beta, rhythmic Spike and Wave (rSW/rPSW), burst suppression, High Amplitude Fast Activity (HAFA) and rhythmic spikes/sharp waves. The results, collectively, point toward more complicated scenarios for seizure generation than simple excessive excitation and insufficient inhibition and suggest that different seizure onset patterns can arise from different cellular and synaptic mechanisms.

## Materials and Methods

### Intracranial electroencephalogram

In a previous work [36], approved by the Centre Hospitalier de l’Université de Montréal (CHUM) Ethics Committee, we reviewed 103 iEEG ictal recordings using intracranial EEG electrodes (128- channel recording system, Harmonie, Stellate, Montreal, Canada, 2000Hz sampling rate) from 20 patients with drug-resistant focal epilepsy who were candidates for resective surgery and, identified nine common seizure onset patterns [7,8]. In the current study, we used some of those seizure onset classes to validate and classify our model simulations.

### Computational model

In the study of epilepsy, the computational approach has been widely utilized in order to provide a mechanistic insight into mechanisms responsible for seizure generation [37,38]. In general, there are two main approaches in constructing mathematical models of ictal activity: biophysically realistic models and phenomenological models. Biophysically realistic modeling approaches rely on detailed cellular and synaptic mechanisms which lead to highly parameterized and complicated models [39–43]. On the other hand, phenomenological models describe the dynamic without reference to mechanisms and are generally more tractable and computationally less expensive [44,45]. These models, while being straightforward in implementation, do not have a physiological equivalent in their elements. Therefore, using phenomenological models, findings are disconnected from physiological phenomena, rendering interpretation more limited when designing therapeutic measures. More recently a hybrid approach has been introduced that combines both biophysically realistic and phenomenological models and is aimed at capturing the benefits of these two approaches through the incorporation of the relevant biophysical features into a simple framework [46].

In this study, we adopted the hybrid modeling approach. We used a well-established coupled neuronal population model, the Jansen model [47], and expanded it with some relevant biophysical features. We incorporated the slow and fast inhibitory mechanisms as suggested by Wendling et al. [35] to respectively account for the dendrite and soma targeting inhibitory interneurons. As presented in Fig.1a, the model consists of four components: the pyramidal cells (PYR), the excitatory interneurons (E), the slow dendrite targeting inhibitory interneurons (I_D_) and the fast soma targeting inhibitory interneurons (I_S_). The postsynaptic activity of the pyramidal cells (the main excitatory component) is considered as the model output to simulate the ictal iEEG recordings.

**Figure 1.**
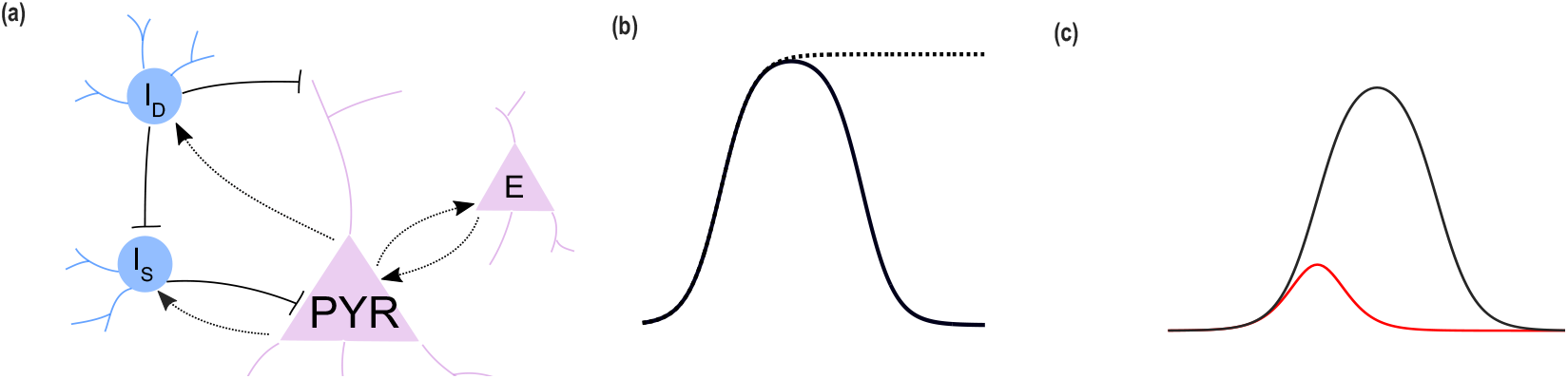
Excitatory inhibitory neuronal populational model and the activation functions. (a) The neuronal population model which is composed of excitatory and inhibitory components: pyramidal cells (PYR), excitatory interneurons (E), dendrite targeting inhibitory interneurons (I_D_) and soma targeting interneurons (I_S_). (b) The nonlinear activation function of a neural population that relates the average postsynaptic potential to an average firing rate for that population. The dotted black curve represents the commonly used sigmoid function and the solid black curve shows the activation function we used which considers depolarization block. (c) Decreasing the depolarization block threshold for the inhibitory populations. In this figure, the red activation function has a lower depolarization block threshold (θ = 0) compared to the black activation function (θ = 4).

In this approach the average postsynaptic potential of each neural population is modeled by two main parts: 1) an activation function that describes a nonlinear relationship between the neural population’s average postsynaptic potentials and its output firing rate and 2) a linear transfer function, 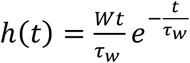, that transforms the input firing rate into average postsynaptic potential. In this equation, W and τ_w_ respectively represent the average synaptic gain and average synaptic time delays for each neural population. The model can be expressed as a system of coupled second-order differential equations [35]:

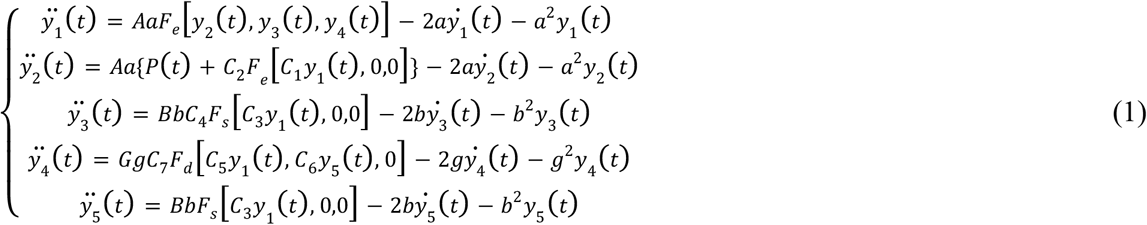

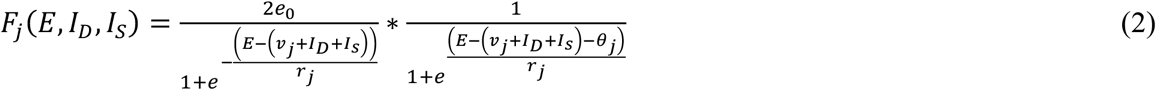

where *A, B* and *G* represent the synaptic gains of respectively the excitatory, dendrite targeting inhibitory interneurons and soma targeting inhibitory interneurons. P(t) describes inputs from other regions which are modeled as Gaussian white noise similar to the original model [47]. Parameters *C*_*1*_ to *C*_*7*_ are the connectivity constants which represent the average number of synaptic contacts (Table 1). In Eq.2, F_j_ where j_∈_ {e,s,d}, represents the activation function for the excitatory population when *j* = *e* and for dendrite and soma targeting inhibitory interneurons when respectively *j* = *d* and *j* = *s*. The variable *θ*_*j*_ represents the depolarization block threshold for dendrite targeting and soma targeting inhibitory interneurons respectively when *j* = *d* and *j* = *s*.

**Table 1.** The model parameters used in Eq.1 and Eq.2.

| Parameter | Value |
| --- | --- |
| $\tau_e$ | 0.01 s |
| $\tau_d$ | 0.02 s |
| $\tau_s$ | 0.002 s |
| $C$ | 135 |
| $C_1, \dots, C_7$ | $C_1 = C, C_2 = 0.8C$<br>$C_3 = C_4 = 0.25C$<br>$C_5 = 0.3C, C_6 = 0.1C$<br>$C_7 = 0.8C$ |
| $e_0$ | $2.5 S^{-1}$ |
| $v_0, r_0$ | 6 mV, 1.7 mV |
| $v_{0i}, r_i$ | 3 mV, 0.6 mV |
| $\theta_e$ | 15 |
| $\theta_s, \theta_d$ | 0 or 4 (Table 2) |

Usually, the activation function is assumed to follow a sigmoid-shaped function; however, more recent studies suggest a bell-shaped activation function which considers depolarization block [48–50]. An activity-driven depolarization block is suggested to be a rate-limiting mechanism to protect cells from excessive firing and can be due to synaptic resources or energy depletion during high levels of activity [49]. Accordingly, in this study, we replaced the commonly used sigmoid function of the original model with the firing rate function in Eq.2 to account for the effect of depolarization block in our model (Fig.1b) [50].

A second mechanism that was incorporated in the model was the collapse and recovery of the inhibitory activity. Some studies have reported seizure initiation due to the failure of inhibition with an important role for the depolarization block of inhibitory cells [28,51–54]. On the other hand, there is experimental evidence for the enhancement of inhibition before and at the very onset of seizure, contradicting the widely accepted paradigm on inhibitory failure mechanisms [29–33,55,56]. To explore both scenarios, we further expanded the model by considering variable depolarization block threshold for the activation function of the inhibitory populations (i.e., *θ*_*s*_, *θ*_*d*_ in Eq.2) (Fig.1c).

In all simulations, the depolarization block threshold of the excitatory populations was set to 15 (i.e., *θ*_*e*_=15) which was chosen to adjust the excitatory activation function to have the same slope at half activation as the sigmoid function used by Jansen et al. [47] in their original model (Table 1). Inhibitory neurons due to their limited size become activated for smaller inputs and reach their depolarization block with a faster dynamic compared to the large pyramidal cells [48]. In order to consider these differences between the excitatory and inhibitory populations in our model, we modified parameters *r*_*i*_ and *v*_*i*_ as presented in Table 1 and chose *θ*_*d*_ and *θ*_*s*_ value to be 4 so that it would have the same slope at half activation as a sigmoid function with parameters *r*_*i*_ and *v*_*i*_. However, wherever we intended to explore the effect of the collapse of an inhibitory population, we reduced the depolarization block threshold of that inhibitory population to zero (i.e., *θ*_*d*_, *θ*_*s*_). Fig.1c depicts the activation function of the inhibitory populations for both depolarization block threshold values equal to 0 and 4 with respectively the red and black curves.

To account for different scenarios of disturbed balance between excitation and inhibition, we used a stochastic approach. In short, the model parameters for excitatory and inhibit synaptic gains (*A, B* and *G*) were divided into one-unit intervals and randomly sampled within each interval. Then, for each interval of the parameters, the most probable pattern of activity was identified. In the next step, starting from the range of *A, B* and *G* parameters, which generated background activity, we examined if the collapse and recovery of the inhibitory activity, which respectively were modeled through decreasing and increasing the parameters *θ*_*d*_ and *θ*_*s*_, could result in the emergence of different seizure onset patterns. For the parameter *A*, we considered a range of values that varied from very low excitation levels to values inducing depolarization block in the excitatory population and explored the effect of different inhibitory modulations in transforming the background activity into ictal patterns; we did not explore “A” values beyond this level.

## Results

We first validated our model by producing some of the commonly observed patterns of ictal activity. Using Eq.1 and Eq.2 for fixed values of *θ*_*e*_, *θ*_*d*_ and *θ*_*s*_ and only by varying the synaptic gains *A, B*, and *G*, we could generate six ictal patterns observed in iEEG recordings as described below (Fig.2):

1. Low Voltage Fast Activity (LVFA): the initial amplitude is lower than 30mV with the frequency range of γ activity (Fig.2(b)),
2. Rhythmic spike/sharp waves: rhythmic spikes with the duration of 20ms to 70ms or sharp waves (duration of 70-200ms); because of the discernable similarity between these two patterns, we merged these two patterns in a single group to simplify our classification (Fig.2(c)),
3. Rhythmic Spike and Wave (rSW): spike and slow wave complexes with the frequency ranging between 2 and 4 Hz (Fig.2(d)),
4. High Amplitude Fast Activity (HAFA): the frequency is greater than 13 Hz and amplitude higher 30mV (Fig.2(e)),
5. Rhythmic α/β: rhythmic activity in the frequency range of α/β activity (Fig.2(f)),
6. Burst suppression: burst of rhythmic spikes interrupted with brief periods of voltage suppression/attenuation (Fig.3(g)).

**Figure 2.**
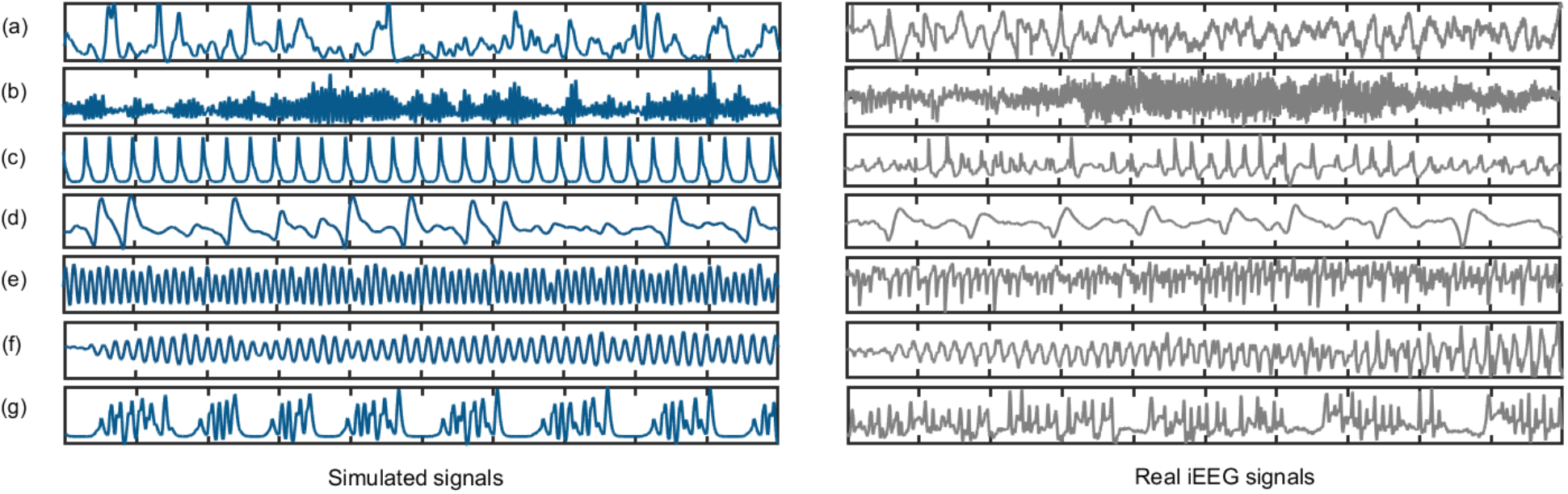
Model simulations vs. real iEEG ictal recordings. The signals in blue are the simulated signals produced by our computational model and the signals in grey are the real depth iEEG recordings for (a) background, (b) Low Voltage Fast Activity (LVFA), (c) rhythmic slow spikes/sharp waves, (d) rhythmic Spike and Wave (rSW), (e) High Amplitude Fast Activity (HAFA), (f) rhythmic α/β and (g) burst suppression ictal activities.

**Figure 3.**
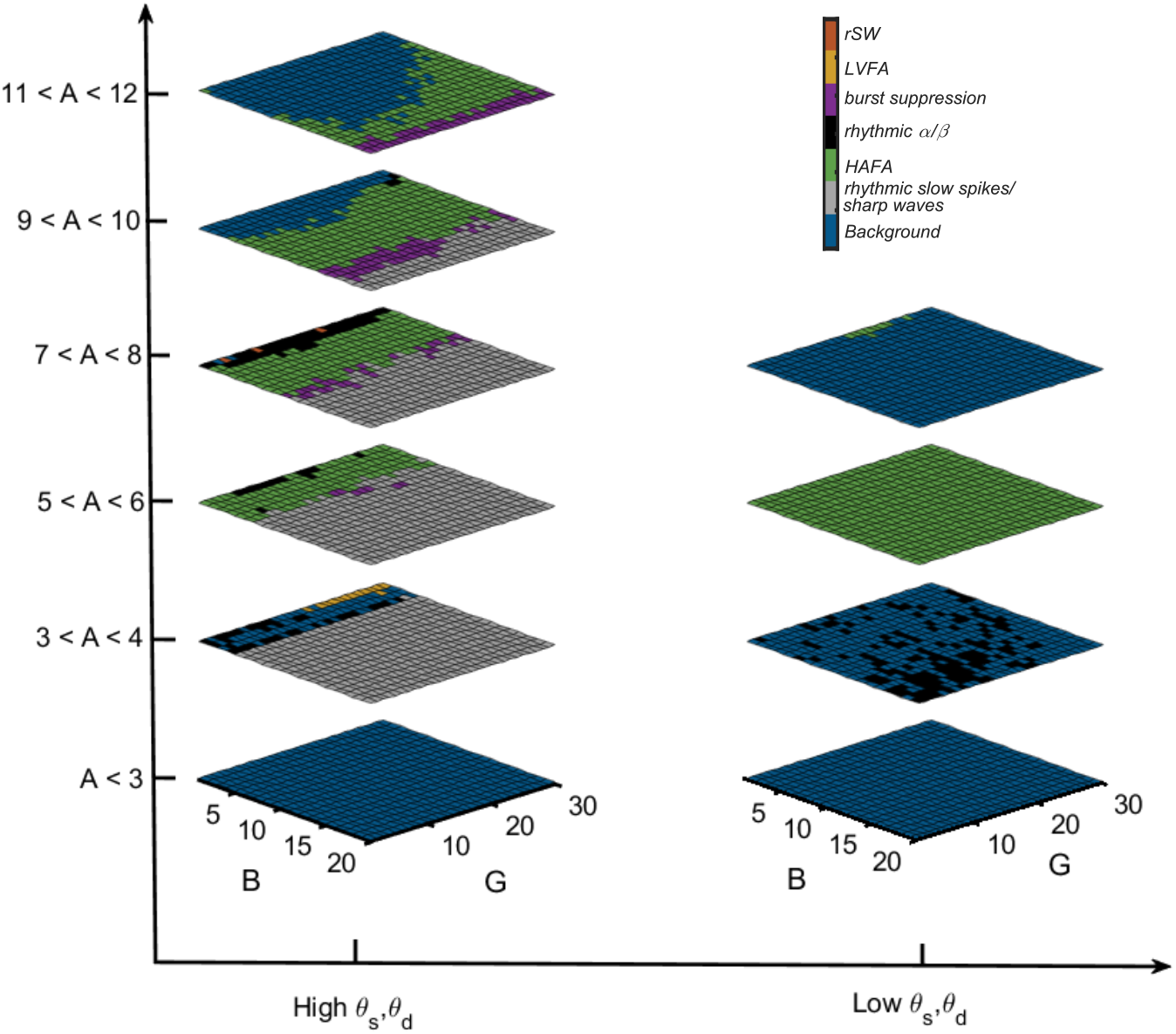
The results from the Monte Carlo simulation. The colors represent the most probable pattern of ictal activity for each one-unit interval of the parameters A, B, and G with θ_d_ = θ_s_ = 4 for the left column and θ_d_ = θ_s_ = 0 for the right column.

To identify the range of parameters producing each activity pattern, we first divided the range of synaptic gains *A, B* and *G* into one-unit intervals. Then, taking the Monte Carlo approach, we drew 100 random values for *A, B* and *G* within each interval and ran the simulations for both low and high values of depolarization block threshold of inhibitory populations (*θ*_*d*_, *θ*_*s*_). In the next step, based on the time and frequency domain features of each simulated signal we classified them into six distinct activity patterns that the model could generate as displayed in Fig.2. We finally identified the range of parameters *A, B* and *G* that generated each pattern of activity for both low and high values of *θ*_*d*_ and *θ*_*s*_ (Fig.3)

The result of our Monte Carlo simulations is presented in Fig.3 and shows that the background activity (shown in blue) was generated for both low and high excitation levels (i.e., *A* values). For the mid-range excitation values, non-background patterns of activity emerge (i.e., ictal activities). However, comparing the left and right columns we can see that for high values of *θ*_*d*_ and *θ*_*s*_, due to higher inhibitory effect, the values of *A* that generate ictal activities (i.e., non-background activity) are less restricted compared to *A* values that produced ictal patterns when low values of *θ*_*d*_ and *θ*_*s*_ are in place.

To explore conditions in which the background activity can turn into an ictal pattern, we started from the background activity (i.e., the blue region in Fig.3) with both low and high *A* values (i.e., low and high levels of excitation) and tested as to whether collapse or recovery of the inhibitory activities (modeled by altering *θ*_*d*_ and *θ*_*s*_) could result in seizure initiation and generation of different seizure onset patterns.

### Seizure generation with low preictal excitation

For values of *A* lower than 3, both low and high values of *θ*_*d*_ and *θ*_*s*_ resulted in background activity –only for 2<A<3 and high *θ*_*d*_ and *θ*_*s*_ for a very narrow range of parameter B was there a low probability of rhythmic slow spikes pattern which was not studied since in our stochastic analysis we only considered the most probable pattern of activity in each interval of the parameters which was the background activity. Therefore, when A<3, by modifying the values of *θ*_*d*_ and *θ*_*s*_ we do not expect to observe a transition from background to ictal activity (i.e., seizure onset).

For *A* values between 3 and 4, the background could be generated for both low and high values of *θ*_*d*_ and *θ*_*s*_. Thus, starting from these two parameter ranges of background activity and by modulating *θ*_*d*_ and *θ*_*s*_, we studied the effect of depolarization block of inhibitory populations in seizure generation. To this end, we fixed *A* to the average value of this interval (3.5) and applying a Monte Carlo approach, found the ranges of *B* and *G* that generated background activities (Fig.4a, Fig.4b). In the next step, for the background activity with high *θ*_*d*_ and *θ*_*s*_ (blue region in Fig.4a), we separately decreased *θ*_*d*_ and *θ*_*s*_ value to 0 and, adopting the Monte Carlo approach, investigated if background activity switched to ictal patterns (i.e., seizures initiated). Similarly, for the background activity with low *θ*_*d*_ and *θ*_*s*_ (Fig.4b), we increased *θ*_*d*_ and *θ*_*s*_ to 4 and explored the possibility of seizure initiation.

**Figure 4.**
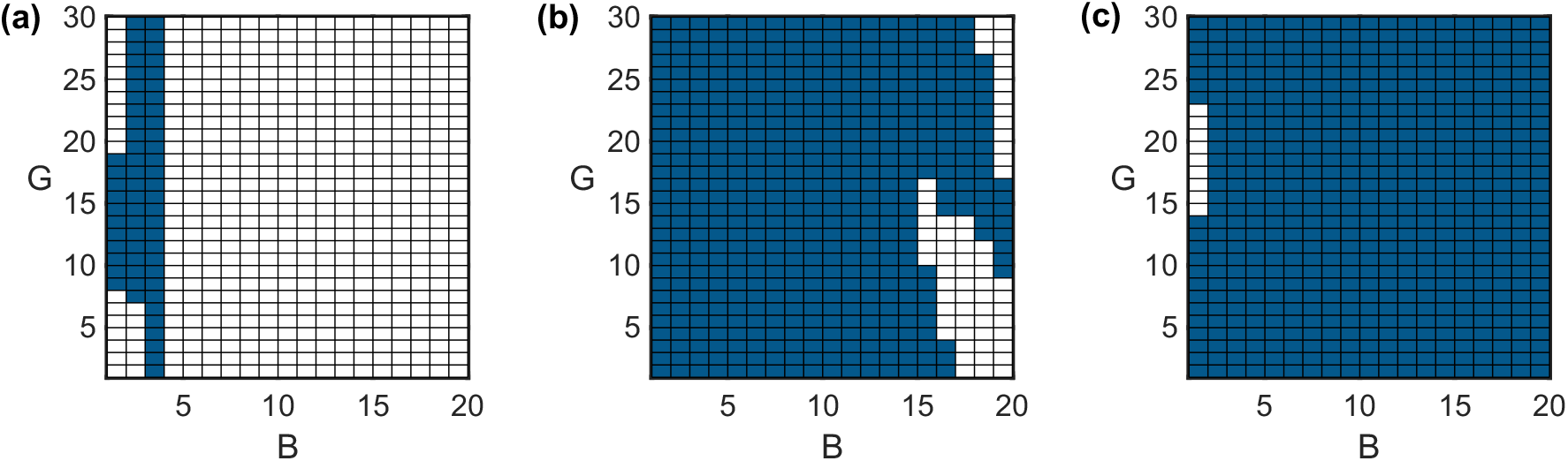
The ranges of parameters B and G generating background activity. (a) For A=3.5 and *θ*_*d*_ = *θ*_*s*_ = 4, (b) for A=3.5 and *θ*_*d*_ = *θ*_*s*_ = 0 (c) for A=7.5 and *θ*_*d*_ = *θ*_*s*_ = 0. The region in blue represents background activity while the region in white corresponds to the range of parameters that did not result in a background activity.

The left column in Fig.5 shows the result of the Monte Carlo simulations for the most probable seizure onset patterns for the first scenario (starting from background activity in Fig.4a). Here, the LVFA seizure onset pattern was generated when only the slow dendrite targeting inhibitory interneurons undergo depolarization block (decreased *θ*_*d*_) which results in disinhibition of fast soma targeting inhibitory activity as well as increased excitation and the subsequent seizure generation. The results for the second scenario are depicted in the right column of Fig.5b where again the LVFA seizure onset is generated mainly by only enhancing the activity of fast soma targeting inhibitory interneuron (increasing *θ*_*s*_ while keeping *θ*_*d*_ = 0). These results are suggestive of the involvement of the fast soma targeting inhibitory interneurons in the generation of LVFA seizure onset pattern when there exist low levels of excitation. These findings are in accordance with the previous modeling and experimental studies reporting this seizure onset pattern when there is high inhibition and low excitation [12,31]. An example of the LVFA seizure onset pattern generated using our model is presented in Fig.6a. For the simulation of this pattern, the parameters *B* and *G* were selected from the ranges specified for this pattern in Fig.5d (Table 2).

**Figure 5.**
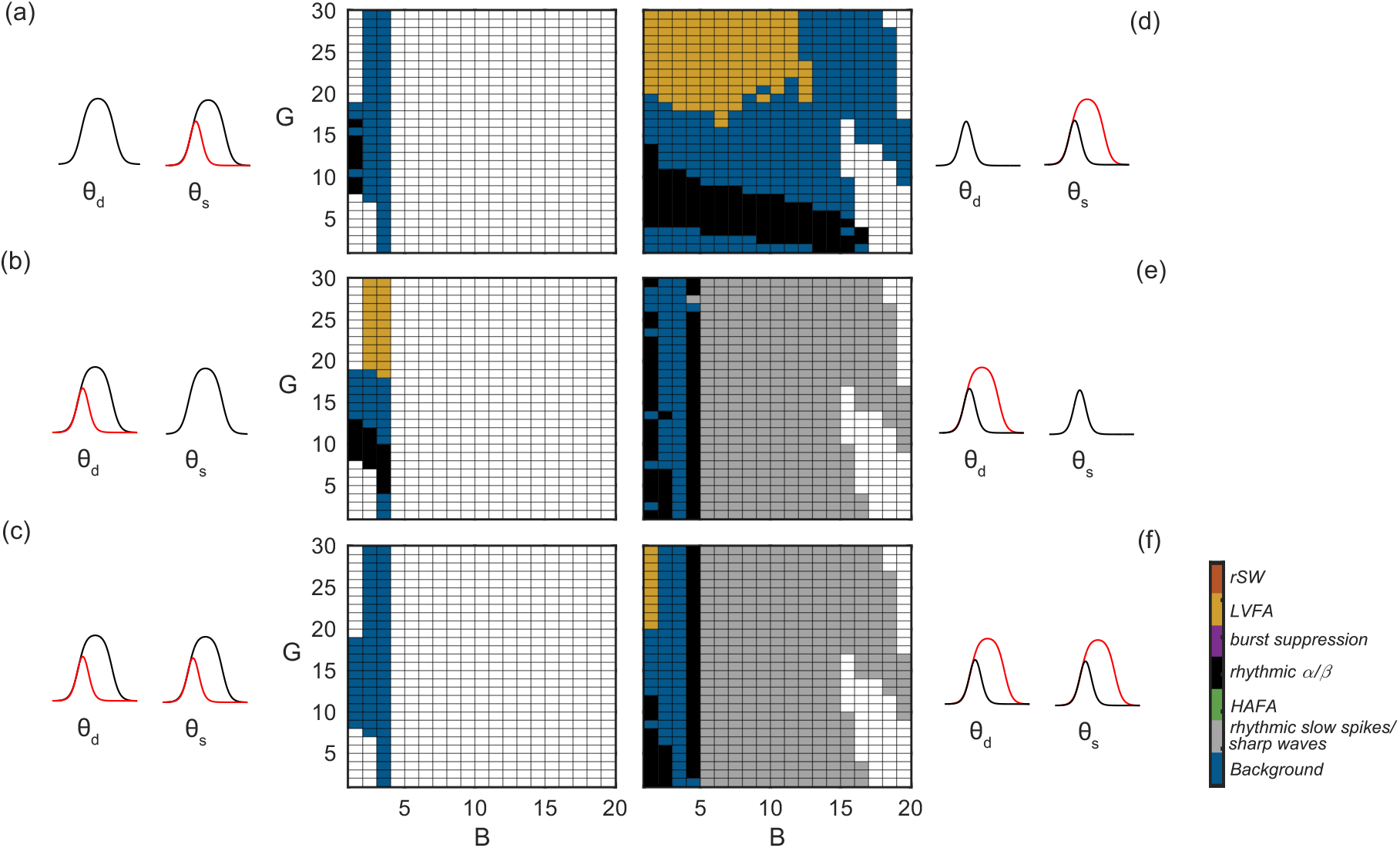
Seizure generation with low preictal excitation. The seizure onset patterns in the left and right columns are respectively generated starting from the background activities represented in Fig.4a and Fig.4b. The underlying mechanism that generated the patterns in the left column is the collapse of (a) soma targeting inhibitory interneurons (*θ*_*s*_: 4 → 0), (b) dendrite targeting inhibitory interneurons (*θ*_*d*_: 4 → 0), (c) both dendrite and soma targeting inhibitory interneurons (*θ*_*d*_: 4 → 0, *θ*_*s*_: 4 → 0) and the mechanism responsible for the generating the onset patterns in the right column is the recovery of (d) soma targeting inhibitory interneurons (*θ*_*s*_: 0 → 4), (e) dendrite targeting inhibitory interneurons (*θ*_*d*_: 0 → 4), (f) both dendrite and soma targeting inhibitory interneurons (*θ*_*d*_: 0 → 4, *θ*_*s*_: 0 → 4). The region in white shows the range of parameters for which there was no background activity to start from and explore the transition to ictal patterns.

**Table 2.** The values used for the simulated patterns. LVFA=Low Voltage Fast Activity, rSW=rhythmic Spike and Wave, HAFA=High Amplitude Fast Activity.

| | Onset pattern | $A$ | $B$ | $G$ | $\theta_d$ | $\theta_s$ |
| --- | --- | --- | --- | --- | --- | --- |
| Fig.6a | LVFA | 3.5 | 7.5 | 28 | 0 | 0→4 |
| Fig.6b | rhythmic $\alpha$ | 3.5 | 1 | 7 | 4→0 | 4 |
| Fig.8a | rSW | 7.5 | 1 | 9 | 0 | 0→4 |
| Fig.8b | rhythmic $\alpha$ | 7.5 | 5 | 15 | 0 | 0→4 |
| Fig.10a | HAFA | 7.5 | 5 | 10 | 0→4 | 0 |
| Fig.10b | burst suppression | 7.5 | 9.2 | 6 | 0→4 | 0 |
| Fig.11a | rhythmic slow spikes | 7.5 | 19 | 20 | 0→4 | 0→4 |

Starting from the background activity with low *θ*_*d*_ and *θ*_*s*_, enhancing *θ*_*d*_ either with or without enhancing *θ*_*s*_, could generate rhythmic spikes/sharp waves onset pattern which shows the importance of slow dendrite targeting inhibitory interneurons for the generation of this onset pattern. Finally, as presented in Fig.5, for a restricted range of parameters *B* and *G*, rhythmic α/β onset patterns are generated (shown in black). Fig.6b presents the onset pattern rhythmic α/β for *B* and *G* selected from the ranges specified for this pattern in Fig.5b (Table 2).

**Figure 6.**
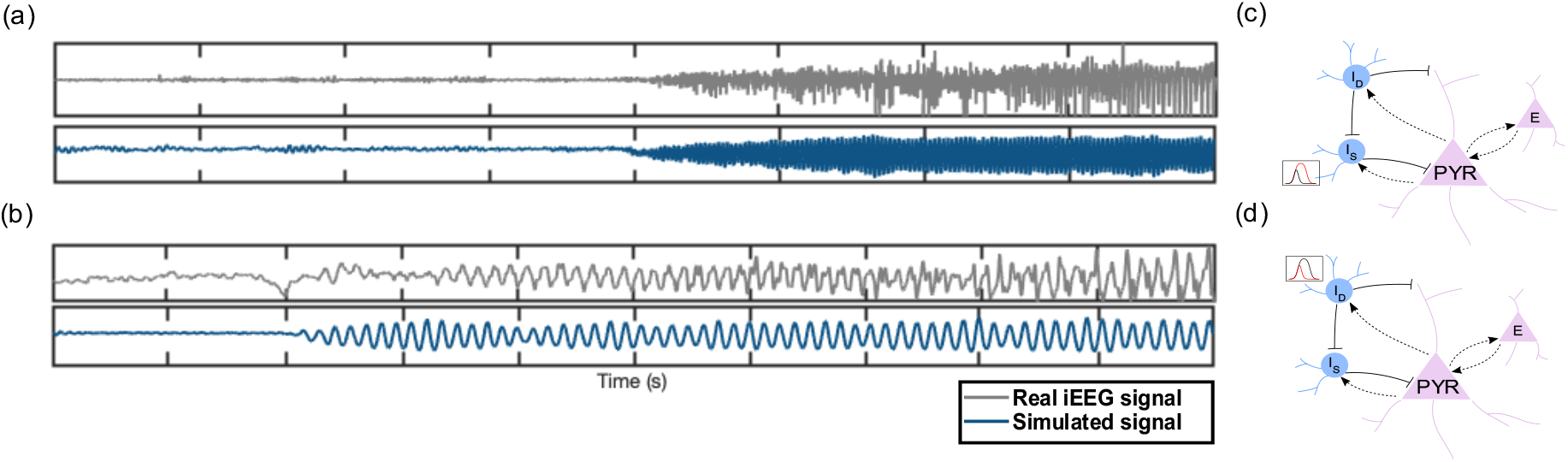
Low Voltage Fast Activity (LVFA) and rhythmic α seizure onset patterns. (a) LVFA seizure onset pattern with low excitation level (i.e., A=3.5). The parameters for the generation of this pattern are chosen from the ranges displayed in Fig.5d and are presented in Table 2. (b) Rhythmic α seizure onset pattern with low excitation level (i.e., A=3.5) and parameters B and G from Fig.5b. (c) The mechanism responsible for the generation of this onset pattern in (a) which is increasing the depolarization block threshold for some targeting interneurons (I_S_). (d) The mechanism responsible for the generation of this onset pattern in (b) which is decreasing the depolarization block threshold for dendrite targeting interneurons (I_D_).

### Seizure generation with high preictal excitation

The other scenario that we considered for seizure generation is the transition from background activity to ictal activity when the excitation level was high (i.e., high values of *A*). The background activity for low *θ*_*d*_ and *θ*_*s*_ occurred when 7<A<8 while for high *θ*_*d*_ and *θ*_*s*_ this range of parameter ‘*A*’ resulted in ictal patterns (Fig.3). Therefore, starting from this background activity with low *θ*_d_ and *θ*_s_, we expected to observe transition to ictal patterns by increasing *θ*_*d*_ and *θ*_*s*_ (recovery of the inhibition). To this end, we fixed *A* to the average value of this range (7.5) and, taking the Monte Carlo approach, identified the range of parameters B and G that generated the background activity (Fig.4c). For this range of parameters (blue region in Fig.4c), we then increased *θ*_*d*_ and *θ*_*s*_ values separately. As presented in Fig.7, increasing only *θ*_*d*_ or both *θ*_*d*_ and *θ*_*s*_ results in seizure generation with mainly three distinct onset patterns: HAFA, burst suppression, and rhythmic spikes/sharp waves (Fig.7b&7c). However, when we only increased *θ*_*s*_ (and not *θ*_*d*_), the seizure onset patterns and the range of parameters generating them were substantially different (Fig.7a). In this scenario, we observed rSW and rhythmic α/β and for a restricted range of parameters HAFA onset patterns. As the dendrite targeting interneurons also inhibits the soma targeting interneurons, high values of *θ*_*d*_ result in suppression of the activity of the soma targeting interneurons, and consequently, changes in parameter G introduce lower variations in the activity patterns we observe (Fig.7b&7c) while low *θ*_*d*_ disinhibits the soma targeting interneurons and allows the parameter *G* to have a more noticeable effect in the patterns that are generated (Fig.7a).

**Figure 7.**
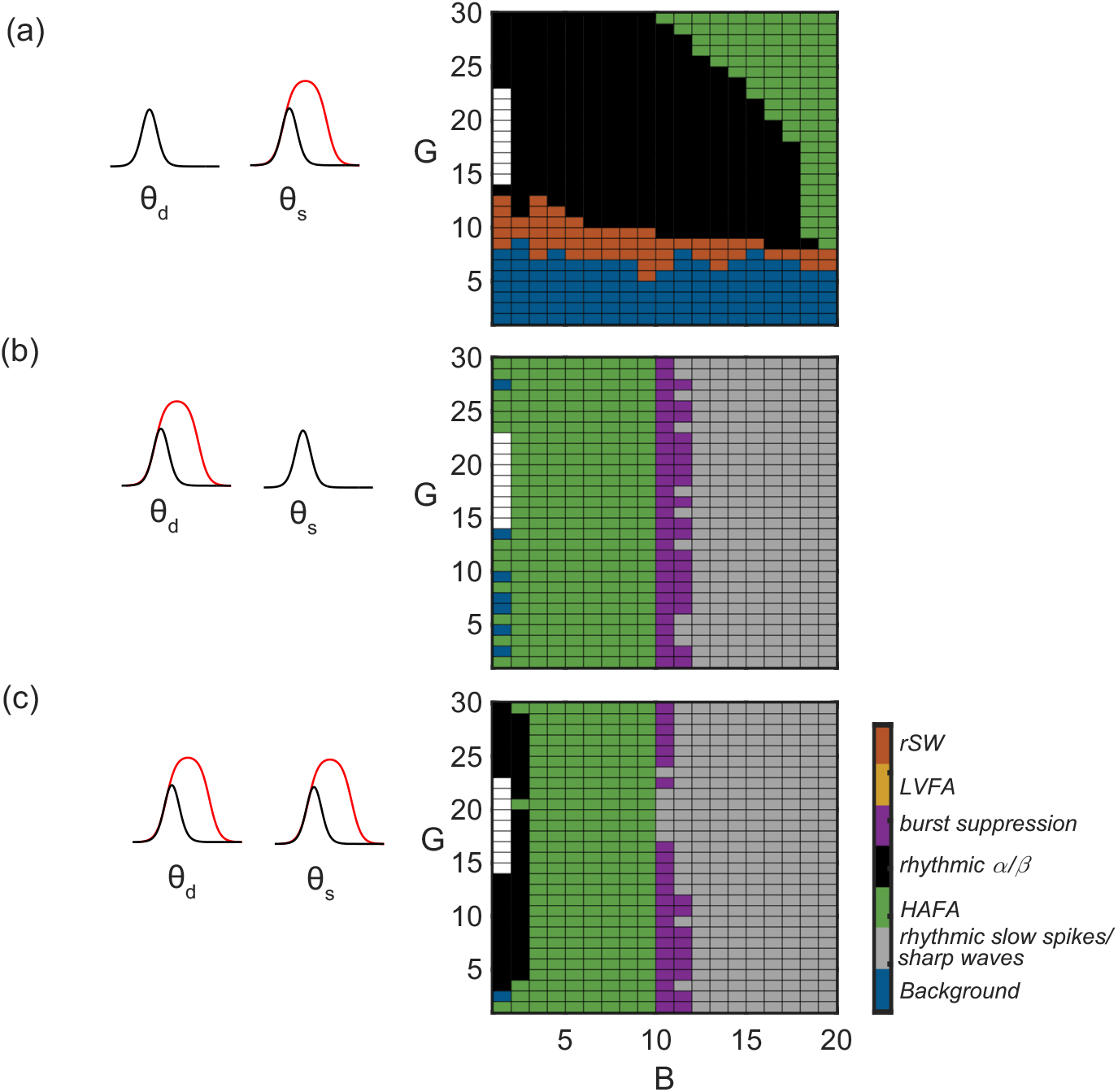
Seizure generation with high preictal excitation. These seizure onset patterns are generated starting from the background activity represented in Fig.4c. The underlying mechanism generating these patterns is the recovery of (a) soma targeting inhibitory interneurons (θ_s_: 0 → 4), (b) dendrite targeting inhibitory interneurons (θ_d_: 0 → 4), (c) both dendrite and soma targeting inhibitory interneurons (θ_d_: 0 → 4, θ_s_: 0 → 4). The region in white shows the range of parameters for which there was no background activity to start from and explore the transition to ictal patterns.

An example of the simulation of rSW seizure onset pattern is presented in Fig.8a. Here, the parameters *B* and *G* were selected from the ranges specified in Fig.7a for rSW onset pattern (Table 2). In this example, enhancing the depolarization block threshold of the soma targeting inhibitory population (*θ*_*s*_) results in recovery of the inhibitory activity which consequently removes the depolarization block of the excitatory population (Fig.8c) and generates rSW seizure onset pattern. Fig.8b shows an example of rhythmic α/β seizure onset pattern. In this example, similar to Fig.8a, the recovery of the soma targeting inhibitory interneurons (*θ*_*s*_) enhances the activity of this population which in turn removes the depolarization block of the excitatory population (Fig.8c). However, here the parameters *B* and *G* were selected from the ranges that generate α/β onset patterns (Fig.7a, Table 2). In Fig.9 as an example, a 3-dimensional representation of the dynamic change in the activation function of the inhibitory population is depicted. Increasing the threshold for depolarization block of the inhibitory population enhances the inhibition (Fig.9a) which in turn draws the excitatory population out of depolarization block (Fig.9b).

**Figure 8.**
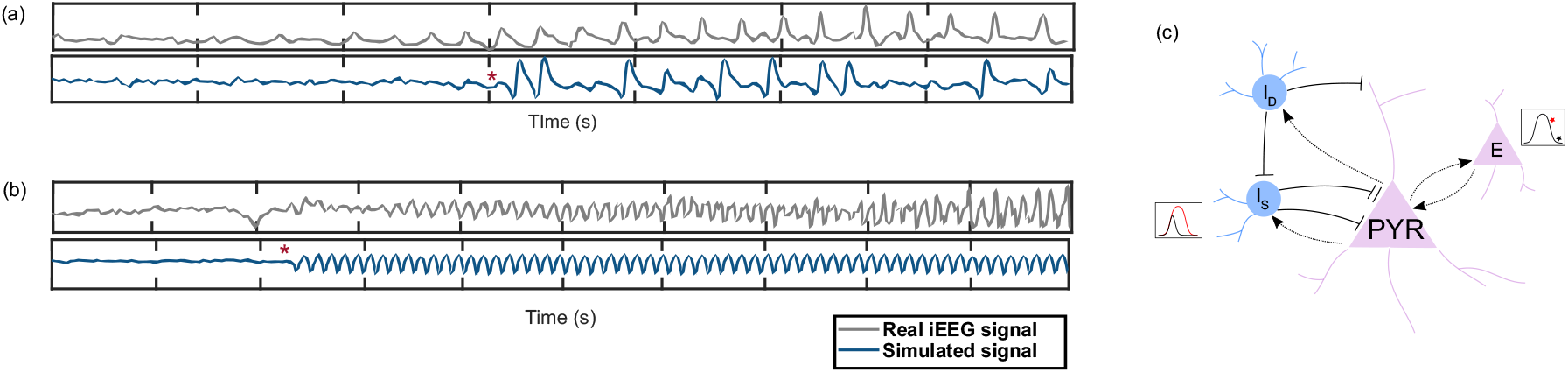
Rhythmic spike and wave (rSW) and rhythmic α seizure onset patterns. (a) RSW and (b) rhythmic α seizure onset patterns generated with the parameters B and G chosen from Fig.7a (Table 2). (c) The mechanism that resulted in the generation of these two patterns in our model which is the recovery of the soma targeting interneurons and the consequent removal of the depolarization block of the excitatory population.

**Figure 9.**
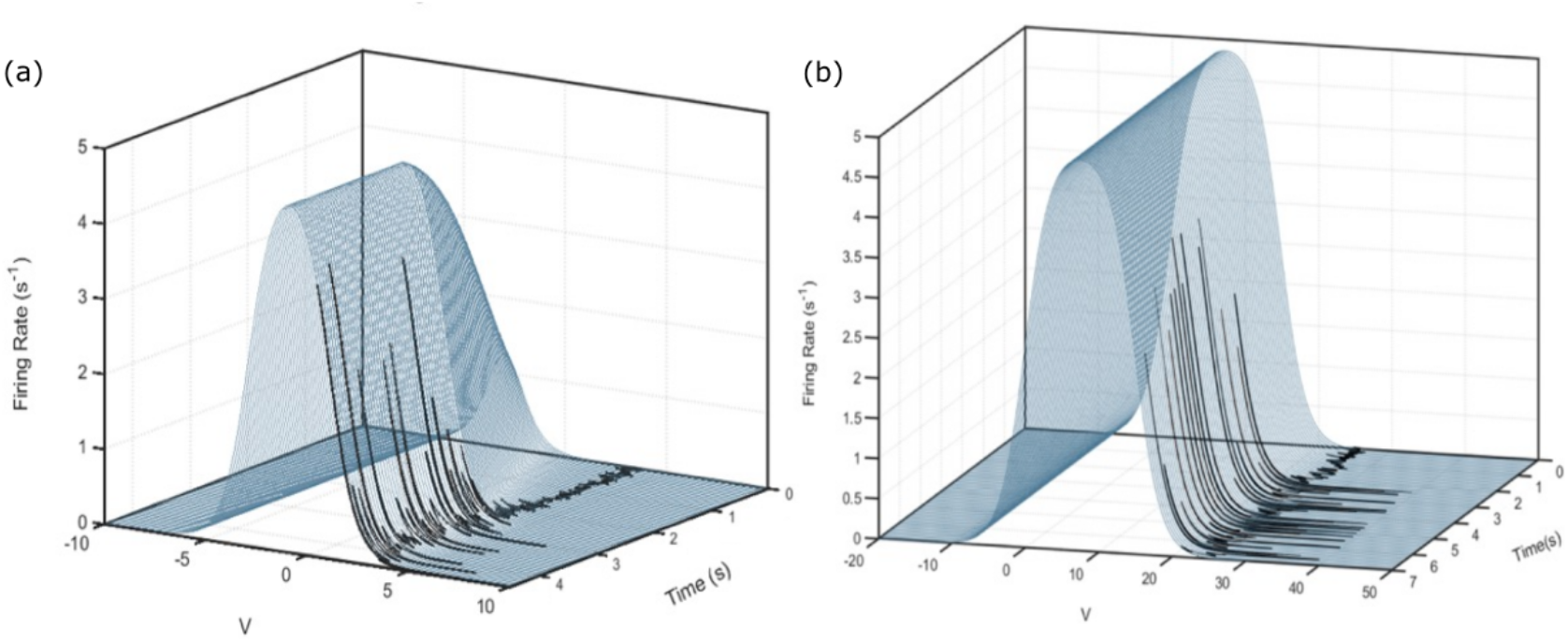
An example for the 3-dimensional representation of the excitatory and inhibitory activation functions. (a) The dynamic change of the inhibitory activation function due to an increased threshold for the depolarization block of this population results in enhanced inhibitory activation. (b) The excitatory population is being drawn out of depolarization block due to its inhibitory input shown in (a).

By the recovery of the inhibitory activity of dendrite targeting interneurons (*θ*_*d*_), Fig.10a and Fig.10b respectively represent an example for HAFA and burst suppression seizure onset patterns with the parameter *B* and *G* selected from the ranges suggested in Fig.7b (Table 2).

**Figure 10.**
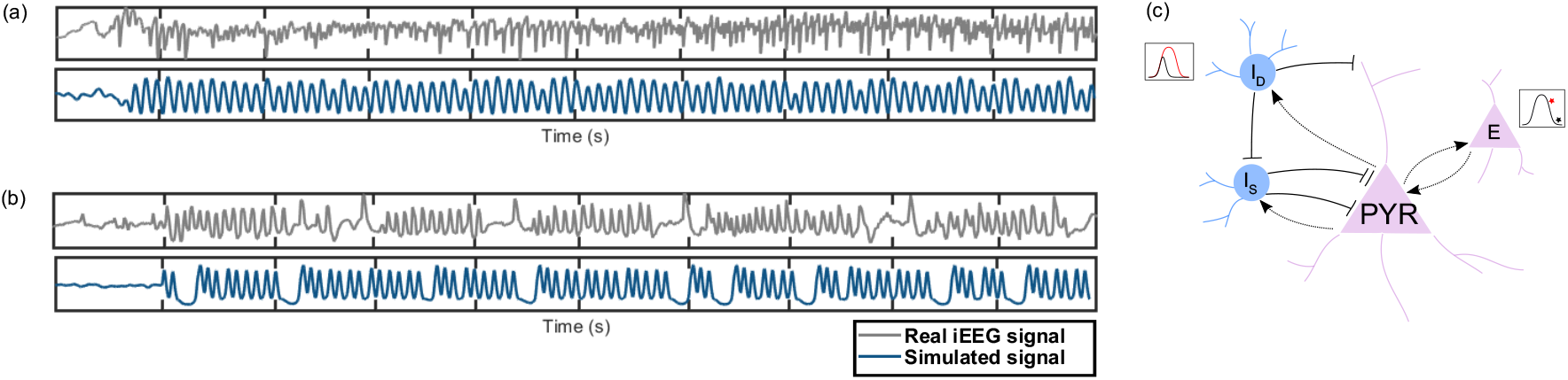
High Amplitude Fast Activity (HAFA) and burst suppression seizure onset patterns. (a) HAFA and (b) burst suppression seizure onset patterns generated with the parameters chosen from Fig.7b (Table 2). The DC shift from the background to ictal activity is removed in this simulated signal. (c) The mechanism responsible for the generation of the onset patterns is shown in (a) and (b). The recovery of dendrite targeting inhibitory interneurons draws the excitatory population out of the depolarization block and initiates seizures.

And finally, by recovery of the inhibitory activities for both dendrite and soma targeting interneurons (*θ*_*d*_ and *θ*_*s*_), Fig.11 depicts an example for rhythmic slow spike seizure onset pattern with the parameter *B* and *G* selected from the ranges suggested in Fig.7c (Table 2).

**Figure 11.**
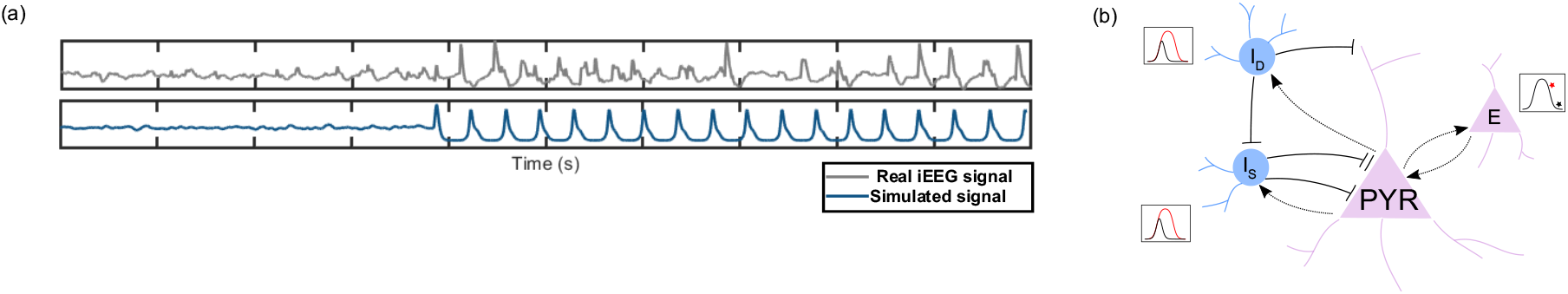
Rhythmic slow spikes seizure onset pattern. (a) Rhythmic slow spikes seizure onset pattern which is generated with the parameters chosen from Fig.7c. The DC shift from the background to ictal activity is removed in this simulated signal. (b) The mechanism resulting in the generation of the onset patterns shown in (a) which is the recovery of both dendrite and soma targeting inhibitory interneurons and the consequent removal of the depolarization block of the excitatory population.

In summary, using our model, LVFA seizure onset pattern could be reproduced mainly by low excitation and high soma targeting inhibitory activity while rSW, HAFA, and burst suppression seizure onset patterns could only be produced when there was high excitation level (high *A*). For the latter due to the high excitatory activity, the excitatory population was experiencing depolarization block and the enhancement of the inhibitory activity removed the depolarization block of the excitatory population and seizures initiated. Two onset patterns - rhythmic α/β and rhythmic spikes/sharp waves - could be generated from the background activity with both high and low excitation levels. When excitation was high, rSW and rhythmic α/β patterns could be generated only when fast soma targeting inhibitory interneurons had recovered. However, the generation of burst suppression and rhythmic spikes/sharp waves onset patterns in this range of parameter ‘*A’* were more related to the recovery of slow dendrite targeting inhibitory interneurons.

## Discussion

We found two main scenarios for the transition from background to ictal activity. Incorporating the effect of depolarization block in our model, the background activity could occur at two different excitation levels: low and high. Under special circumstances, the background activity with both these excitation levels could transition into ictal activity (i.e., seizure generation). Depending on the excitation level, our results showed that different inhibitory modulatory mechanisms - which will be further discussed below- could transform the background into ictal activity. These findings could provide an explanation for the experimental observations that report different behavior of the excitatory populations when they are in different excitability states (e.g., [57]). The optogenetic stimulation of excitatory populations has been reported to induced seizure-like activity when the membrane potential is close to the resting level (i.e., transition from background activity to ictal events) while the photostimulation of the same excitatory neural populations during an ongoing seizure-like-event terminate ictal discharges (i.e., return to the background activity) [57]. On a larger scale as well, the emergence of the background activity due to high excitation has been observed when the application of deep brain stimulation suppresses ongoing seizures (e.g., [58]). Depolarization block mechanism can offer an explanation for this dual effect of neural stimulation in seizure generation and termination and in general, could suggest pathways for seizure generation and termination mechanism. As presented in the schematic Fig.12, this bell-shaped activation function suggests that increasing the excitation level (e.g., by the application of electrical stimulation), depending on the current state of the excitatory population, can result in seizure initiation (Fig.12, Onset I) or termination (Fig.12, Termination I). Similarly, based on the current state of the excitatory population, decreasing the excitation level (e.g., by enhancing inhibitory activity) can convert the background activity into ictal patterns (i.e., initiate seizures) (Fig.12, Onset II) or terminate seizures (Fig.12, Termination II).

**Figure 12.**
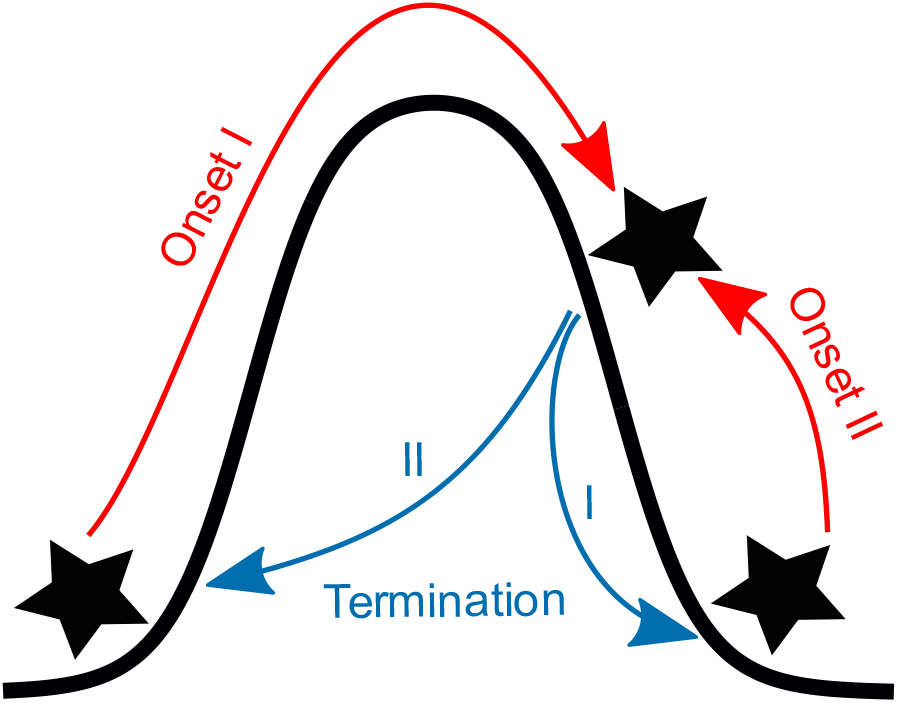
A schematic figure depicting our suggested seizure onset and termination pathways considering the depolarization block mechanism. Depending on the initial excitability state, enhancing the excitation level could result in both seizure generation (Onset I) and termination (Termination I), similarly, decreasing the excitation level could result in both seizure generation (Onset II) and termination (Termination II).

Our model showed that in the presence of different excitation levels and specific range of synaptic gain parameters both the enhancement and collapse of inhibitory activity could result in the generation of seizures. These findings could unify the seemingly contradictory mechanisms suggested by previous experimental studies for the involvement of inhibitory activity in seizure generation [25–33,55]. While the collapse of inhibition has been widely reported as a main mechanism underlying seizure generation [25–28], some experimental studies have addressed the enhanced inhibitory activity before and at the very onset of ictal activities [29–33,55,56]. In the current study, we found that depending on the current excitation level, both scenarios could result in the generation of seizures. More specifically, with low levels of excitation both the collapse and enhancement of inhibition could initiate seizures with different onset patterns while with high levels of excitation, only the enhancement of inhibitory activity could remove the depolarization block of the excitatory population and initiate seizures. Therefore, based on our modeling approach, depending on the excitation level and the range of synaptic gains, both collapse and enhancement of inhibitory activities could transform the background activity into ictal patterns (i.e., initiate seizures).

Our results point towards different underlying mechanisms for the generation of seizures with different onset patterns. For instance, seizures with LVFA onset pattern could be generated when there was low level of excitation in the model and enhanced somatic inhibition either due to the recovery of this inhibitory population or because of being disinhibited due to the collapse of dendrite targeting interneurons. These findings are in agreement with experimental studies suggesting enhanced inhibitory activity and low excitation at the onset of seizures with this onset pattern [12,18,30,31]. In the well-known framework [35] based on which we constructed our model, the impaired dendrite inhibition is reported responsible for the generation of LVFA onset pattern. Similar to our framework, in their model, the collapse of dendrite targeting interneurons which was considered as the direct mechanism generating this onset pattern, results in disinhibition of the soma targeting interneurons which could be an indirect mechanism responsible for the generation of this seizure onset pattern [35]. The repetitive spiking seizure onset pattern which is mainly attributed to seizures with high amplitude periodic spikes [59–61] as well as the bursting events, are reported to occur when the excitation is high [62,63] which agrees with our model findings. Our results showed that HAFA, rSW and burst suppression seizure onset patterns could be reproduced due to the recovery of the inhibitory activity only when the excitation level was high, and the excitatory population was experiencing depolarization block. The recovery of the inhibition removed the depolarization block of the excitatory population and initiated seizures.

Taken together, in order to explore the underlying mechanisms for the generation of seizures with different onset patterns, we constructed an excitatory-inhibitory neural population model and considered depolarization block mechanism and modulated the modeled slow and fast inhibitory activities. By including the depolarization block mechanism, our results showed that the background activity is possible for both low and high excitation levels (i.e., *A* values). Starting from these two background activities, we examined different scenarios in which a background state could transition into ictal patterns and explored whether seizures could be generated by modulating the inhibitory activities. Our results showed that for specific range of parameters both the enhancement and collapse of inhibitory activity could result in seizure initiation. By reproducing some of the most commonly observed seizure onset patterns, our findings suggest that the mechanisms responsible for seizure generation are different in seizures with distinct onset patterns. Understanding these different seizure generation mechanisms could aid us towards better seizure control measures. While, in this study, we mainly considered the modulation of the activation function of the inhibitory populations (the collapse and recovery of the inhibition), an interesting expansion of this model could be incorporating the modulatory effect of the inhibitory population on the activation function of the excitatory population which could happen for instance due to shunting inhibition and GABAergic depolarization.

## Data Availability

All data produced in the present study are available upon reasonable request to the authors

## Acknowledgements

This research was supported by CIHR project grant (390044); Canada Research Chair Program (DKN).

